# Task-oriented vs. Goal-oriented: A New Paradigm for “Rooming” Patients

**DOI:** 10.1101/2024.10.17.24315639

**Authors:** Temesgen Tsige, Matthew Bacchus, Anthony Flora, Gabriella Ahn, Sandhya LoGalbo, Vinayak Sahadeo, Prajna Sahadeo, Mark Richman

## Abstract

**INTRODUCTION:** In the fast-paced setting of the Emergency Department (ED), efficiency is important to delivering timely, effective care. A key aspect of this involves preparing patients by providing hospital gowns and connecting them to cardiac/vital signs monitors. These seemingly basic steps are crucial for ensuring patient safety and streamlining the provider’s evaluation process. However, in the ED, patients are often not fully prepared before a provider enters the room, leading to delays and inefficiencies. This study examines the rates at which patients are properly gowned and monitored upon rooming, to identify areas where ED workflows can be optimized to improve care delivery and better utilize staff and resources.

**MATERIALS AND METHODS:** The study was conducted in the adult Emergency Department at Long Island Jewish Medical Center. Data were collected from 100 randomly selected patients between June and October 2024. Patients were excluded if they wore loose-fitting clothing that allowed easy access to the area being evaluated, or if their chief complaint involved a visibly accessible body part (e.g., face), eliminating the need for disrobing or monitoring. The need for a hospital gown or connection to a cardiac/vital signs monitor was determined based on standard triage criteria, such as cardiopulmonary symptoms or the need to examine areas not easily accessible through clothing. We measured the percentage of patients who were appropriately gowned or monitored prior to the provider entering the room.

**RESULTS:** Of the 100 patients, 89 required a hospital gown, but only 61 (69%) received one. Regarding cardiac monitoring, 46 patients required a monitor, but only 20 (43%) were connected to one. In 50% of cases, patients had the correct body part exposed for examination before the provider’s arrival.

**CONCLUSION:** This study highlights significant inefficiencies in patient preparation in the ED, specifically related to gowning and cardiac monitoring. While 89% of patients required a gown, only 69% had one before the provider’s arrival. Similarly, only 43% of the patients who required monitoring were appropriately connected to a monitor. Additionally, only half of the patients had the correct body parts exposed for examination. These gaps in preparation force providers to complete these tasks themselves, contributing to delays, increasing provider workload, and potentially leading to burnout. To address this, we recommend shifting from task-oriented to goal-oriented patient rooming, revising protocols, and implementing checklists. These changes could help optimize resource use, reduce delays, and improve overall efficiency.

## INTRODUCTION

In the fast-paced and high-stakes environment of the Emergency Department (ED), maximizing efficiency is crucial for ensuring timely and appropriate care^2^. Simple, yet important, steps, such as providing a hospital gown or connecting a patient to a cardiac/vital signs monitor, form the foundation of proper patient preparation. Hospital gowns serve multiple essential purposes. First, they serve as a protective barrier for patients and healthcare workers, safeguarding against exposure to blood and other infectious materials^8^. Second, gowns facilitate clinical assessment by allowing easy access to the relevant areas of the body that need to be examined or treated^11^. Likewise, for patients presenting with cardiopulmonary complaints, the immediate use of a cardiac/vital signs monitor is crucial to track vital signs and identify life-threatening conditions early^17^. Poor preparation prior to the first provider-patient assessment may affect the timeliness of care.

Different health systems assign various staff members - nurses, ED Technicians (repurposed paramedics), patient care associates (PCAs), and medical assistants - to perform essential triage and rooming tasks, including providing necessary personal protective equipment (such as gowns) and cardiac/vital signs monitors, based on the chief complaint and information gleaned during triage. Unfortunately, we have observed during our experience of thousands of hours in the ED that patients often do not have these basic requirements (provision of gown and monitor) met before the provider enters the examination room. Consequently, providers expend time to help dress patients; place ECG leads, blood pressure cuff, and pulse oximeter; and start the cardiac monitor. Failure to comprehensively prepare patients for their face-to-face provider evaluation is a suboptimal use of time and resources for providers, patients, and hospital institutions, particularly as providers are the highest-paid members of the care team^12^. Practicing below their limit of one’s license is a common reason for provider dissatisfaction and burnout^4^.

One framework that might explain the failure to have a patient not only roomed, but also gowned and on a monitor is a “goal-oriented” vs. “task-oriented” mindset^14^. Goal-oriented people utilize a broader temporal frame of reference and anticipate future events, whereas task-oriented people focus on incremental objectives. Therefore, a task-oriented person who views their role in “rooming” a patient solely as the incremental step of moving a patient from the ED waiting room to an examination room is likely not thinking of the next step in the patient’s care: the provider being able to expeditiously and thoroughly evaluate the patient, and the patient’s continued safety through cardiac/vital sign monitoring. Although task-oriented methods may initially seem quicker, they can lead to significant delays and drawbacks, including higher rates of avoidable errors and accidents resulting from inadequate care quality. Ultimately, this undermines the effectiveness and safety of patient care^1^.

ED overcrowding further worsens this issue. High patient volumes strain resources and force healthcare providers to balance speed with accuracy, often at the expense of thorough preparation. Overcrowding is a well-documented phenomenon linked to higher mortality and morbidity rates, as well as decreased quality of care^9,10,13,15,16^. To mitigate overcrowding, EDs typically focus on three strategies: reducing patient input, optimizing patient throughput, and increasing output^3^. While these strategies are essential, they are often applied in a task-oriented manner that prioritizes the movement of patients over the readiness of patients for care. As a result, critical steps, such as providing a gown or connecting a vital sign/cardiac monitor, are sometimes missed or delayed, leading to suboptimal patient experiences and increased workload for providers.

This investigation aimed to understand the rates of appropriate patient gowning and monitoring shortly after a patient is “roomed.” Such information can be used to encourage ED administrators to modify policies, procedures, and expectations so patients can receive more timely and appropriate care and the ED can have a more-optimal use of staff resources, including having all staff practice to the level of their license. This investigation might encourage leadership at other institutions’ ED to investigate their processes of “rooming” patients.

## MATERIALS AND METHODS

This study was conducted at Long Island Jewish Medical Center (LIJ), part of the Northwell Health system, which includes 23 hospitals and 890 outpatient facilities across New York. LIJ is a 583-bed tertiary-care academic hospital that provides care to a diverse population across a range of racial and socioeconomic backgrounds. The study focused on patients admitted to the LIJ adult ED, which handles approximately 100,000 patient visits annually.

The study population included patients admitted to the LIJMC Adult ED. Patients arrive at the ED either by ambulance or by walking into the ED and are escorted to examination rooms by a nurse, PCA, or ED Technician. Patients were identified on the ED tracking board as having been placed in an examination room and not having yet been seen by a provider. Patients were excluded from the study if they wore loose-fitting clothing that could easily be adjusted to expose the necessary body part for evaluation or if their chief complaint involved an easily-visible area (eg, face) that did not require disrobing or the use of a cardiac/vital signs monitor. Patients needing immediate attention, for whom there was essentially no time delay between “rooming” and when the provider arrived, were also excluded. After a 5 minute window following being placed in an examination room (to give the patient an opportunity to change into a gown or be placed on a cardiac/vital signs monitor), a provider and a research assistant entered the room and witnessed whether the patient was appropriately exposed for the intended physical examination and, if needed, was on a monitor.

Patients were considered to require a hospital gown and/or cardiac/vital signs monitor what would typically be expected following a brief triage process. For example, patients with cardiopulmonary symptoms such as chest pain, shortness of breath, or palpitations were expected to be connected to a cardiac/vital signs monitor. Similarly, patients with complaints requiring a physical examination of body parts not easily accessible through clothing (eg, the abdomen, back, or chest) were expected to be in a gown for proper examination.

Data collection was between June and October 2024. Observations were recorded regarding the need for a hospital gown, whether the patient was in a gown, the need for a cardiac/vital signs monitor, and whether the patient was being monitored with such monitor. For each patient, responses were recorded as “yes” or “no.” In cases in which a gown or cardiac/vital signs monitor was not applicable, an “N/A” response was recorded for the monitor variables.

We evaluated 100 randomly-selected patients and calculated the percentage who were in a gown or on a cardiac/vital signs monitor prior to the provider entering the examination room when it was necessary for their clinical evaluation.

This project was reviewed and deemed not to meet the definition of research by the Northwell Health Institutional Review Board’s (IRB’s) Human Research Protection Program (HSRD24-0133), indicating that formal IRB approval was not required for this study. All observations were collected in compliance with institutional guidelines for patient privacy and data security.

## RESULTS

Of the 100 patients, 89 (89%) needed gowns, but only 61 patients (69% of the 89 needing gowns) received them. For cardiac monitoring, 46 patients (46%) needed a cardiac monitor, but only 20 patients (43%) had one (**Figure 1**). Additionally, proper body part exposure during examinations was achieved for only 50 patients (50%) **(Figure 2).**

**Figure 1:**
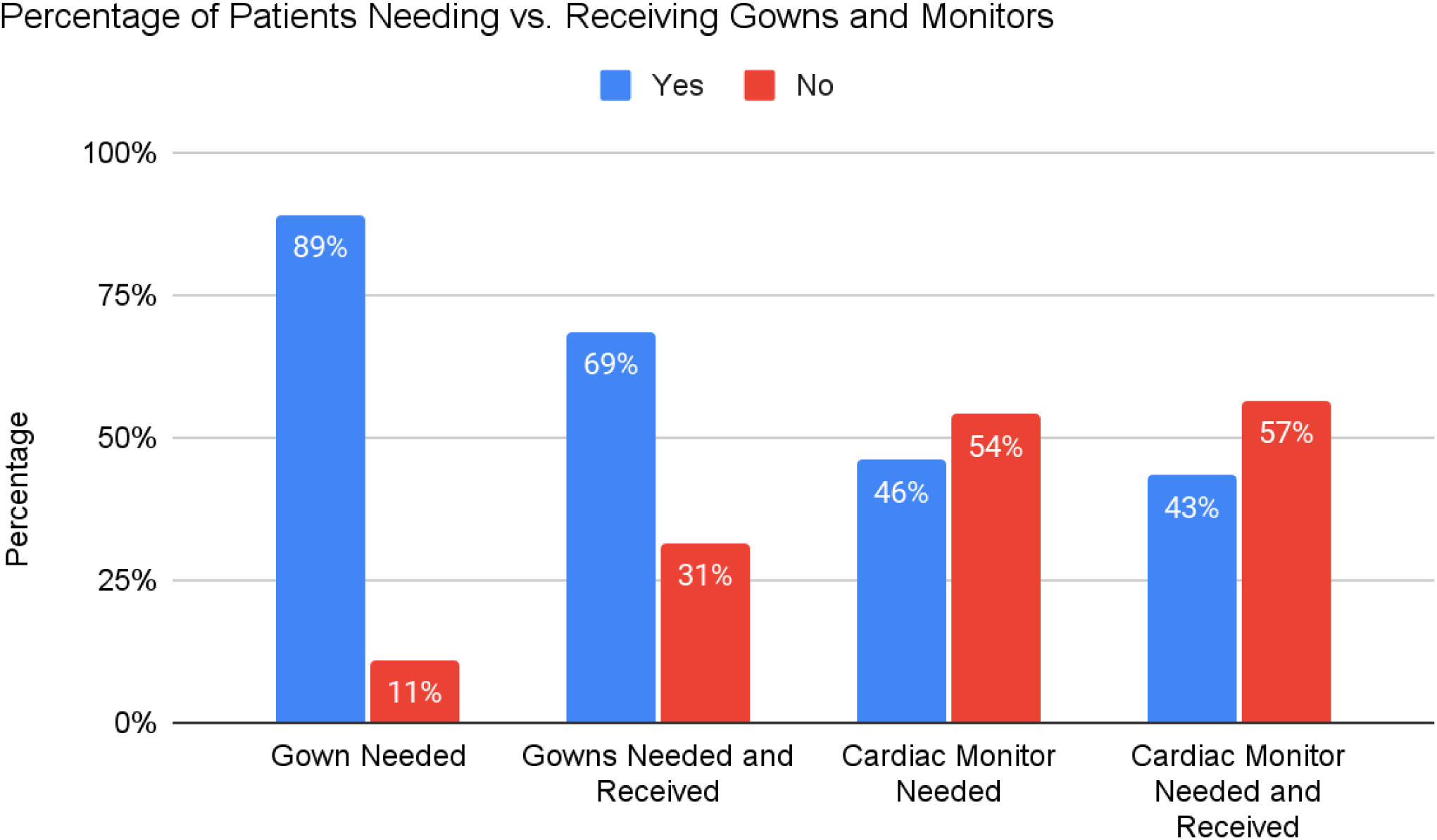
Percentage of Patients Needing vs. Receiving Gowns and Cardiac Monitors

**Figure 2:**
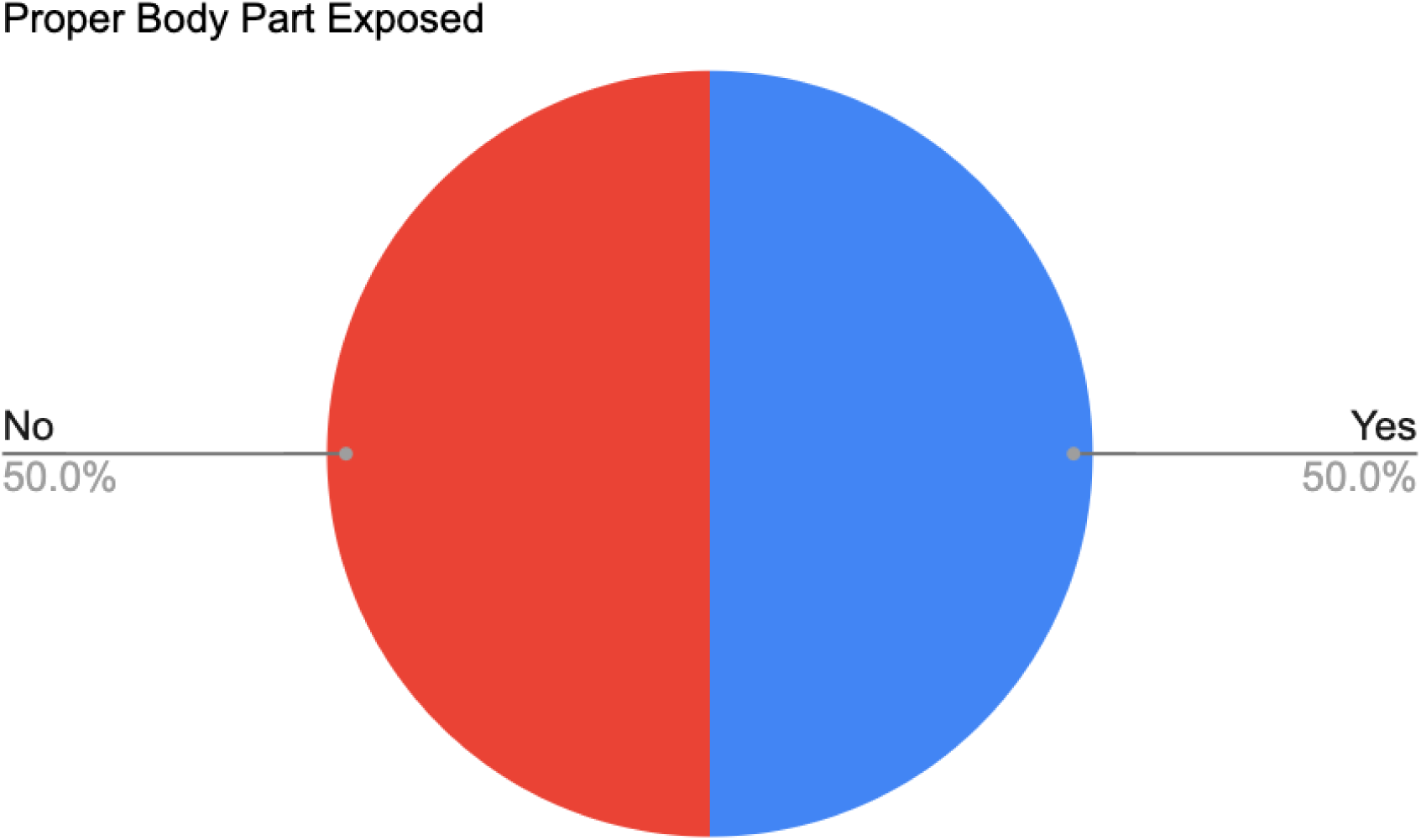
Exposure of Proper Body Parts During Patient Examination

## DISCUSSION

Our study identified inefficiencies in patient preparation regarding gowns and cardiac/vital signs monitors used in the ED. Eighty-nine percent of patients needed a gown, but only 69% of those who required one had it provided before the provider arrived in the patient room. Forty-six percent of patients required a monitor, but only 43% of those had one in place when the provider arrived. Additionally, only 50% of patients had the appropriate body parts exposed for examination, which suggests inefficiencies in ensuring proper exam readiness. It’s important to clarify that while some chief complaints don’t require hospital gowns, they do need proper exposure. For example, a foot complaint doesn’t need a gown, but you must have the foot unobstructed for examination. This allows the healthcare provider to accurately assess any injuries, swelling, or underlying conditions.

Providers entering examination rooms where basic preparation tasks have not been completed must take on roles that are typically not expected of providers, such as dressing patients or setting up monitors. This diversion of responsibilities wastes valuable provider time, limits their capacity to handle more patients, and may contribute to provider burnout^4^. The situation is especially concerning in high-pressure environments such as the ED, where timely completion of tasks can significantly affect patient outcomes^2,6^.

Failure to provide hospital gowns and cardiac/vital sign monitors when needed may be due to various factors. One is the reliance on task-oriented behavior, in which staff prioritizes “rooming” the patient without ensuring readiness for provider evaluation. This approach may appear efficient in terms of moving patients through the ED, but it ultimately delays delivery of care. Second, there may be a misunderstanding (by ED leadership or front-line staff) about the applicability of “goal-oriented” vs. “task-oriented” behavior in “rooming” the patient. Finally, the provider is ultimately the healthcare worker for whom the patient presents to the ED who is responsible for the patient. Upstream healthcare workers may feel confident that, if the patient has not been adequately prepared for evaluation and monitoring, the provider will do so to ensure appropriate management and safety.

To enhance care delivery, institutions must consider their care philosophy, management approach, and available resources. The conceptual framework that informs decision-making is important in shaping how models or policies are selected and implemented. By adopting a “goal-oriented” approach, healthcare organizations can overcome the limitations of task-oriented practices (such as by having patients fully prepared for provider evaluation, rather than just “roomed”), ultimately leading to more thorough and effective patient care^1^.

In addition, overcrowding in the ED worsens these inefficiencies. With high patient volumes and limited resources, staff upstream to the provider may feel pressured to rush through triage and preparation, leaving critical tasks unfinished^5,7^. This can create a bottleneck effect where providers are forced to perform basic tasks that could have been handled earlier by other members of the healthcare team^4^. The random selection of patients ensured patients were triaged and taken to a room by a wide variety of nurses, PCAs, and ED Technicians. Hence, this issue is not simply one of individual negligence, as, but, rather a systemic problem..

To address the inefficiencies identified in this study, ED leadership should consider revising policies and protocols related to patient preparation. For example, whoever is responsible for “rooming” the patient might also be responsible for ensuring the provider can quickly examine the affected body part and that the patient is monitored, if needed. Such a policy might include: 1) Any patient with a pelvic complaint should be undressed to the undergarments; 2) Any patient with an extremity-related chief complaint (eg, pain, injury, rash) should have the affected part exposed; 3) Any patient with a cardiopulmonary complaint should be on a cardiac/vital signs monitor; etc. Implementing checklists or other tools to confirm that each patient is properly prepared could help bridge the gap between task completion and goal achievement^18^.

By improving patient readiness, the ED can optimize the use of staff resources, reduce delays in care, and alleviate some of the burden on providers. In doing so, hospitals may see improvements in patient outcomes, overall workflow efficiency, and provider satisfaction.

## Limitations

This study has several limitations. First, it was conducted at a single center, which may limit the generalizability of the findings to other institutions. Variations in clinical protocols - such as differences in the role of healthcare workers and how protocols are implemented, along with resource availability across different settings - could lead to differing results. Confounding variables may also affect study interpretation. For instance, all patients placed on monitors might have been aged 65 and above. Furthermore, differences in the severity and history of cardiovascular events or chronic conditions might have influenced whether patients received gowns and monitoring. However, random patient selection for the study makes such confounders unlikely. And data do not support such a possibility. For example, patients requiring a monitor but who didn’t receive one included older patients with chest pain and shortness of breath. Additionally, variations in wait times between patient room placement and data collection could impact results; some patients waited 30 minutes while others waited only 5 minutes. Standardizing data collection to a specific time frame after room placement, such as within 5 minutes, could yield different findings. However, as none of the patients was seen by a provider prior to 5 minutes after being “roomed,” and many patients were still without a gown or monitor long after that, this variation is unlikely to affect the interpretation of results. Finally, as an exploratory study to determine the scope of the problem, only a univariate analysis was performed.

## Data Availability

All data produced in the present study are available upon reasonable request to the authors

## Author Contributions

Mark Richman, Matthew Bacchus, Gabriella Ahn, Sandhya LoGalbo, Vinayak Sahadeo: Conceived of the project and did the initial manuscript draft and literature search.

Mark Richman, Prajna Sahadeo, Anthony Flora, Temesgen Tsige, Vinayak Sahadeo, Gabriella Ahn: Created data collection tool, performed data collection and entry, integrated those findings and references into the manuscript, reviewed and edited final manuscript.

## Patient’s Consent

N/A

## Trial/Systematic Review Registry

N/A

## Acknowledgements

None

